# Biomedical Text Normalization through Generative Modeling

**DOI:** 10.1101/2024.09.30.24314663

**Authors:** Jacob S. Berkowitz, Apoorva Srinivasan, Jose Miguel Acitores Cortina, Yasaman Fatapour, Nicholas P Tatonetti

## Abstract

**Objective:** Around 80% of electronic health record (EHR) data consists of unstructured medical language text. The formatting of this text is often flexible and inconsistent, making it challenging to use for predictive modeling, clinical decision support, and data mining. Large language models’ (LLMs) ability understand context and semantic variations makes them promising tools for standardizing medical text. In this study, we develop and assess clinical text normalization pipelines built using large-language models.

**Methods:** We implemented four LLM-based normalization strategies (Zero-Shot Recall, Prompt Recall, Semantic Search, and Retrieval-Augmented Generation based normalization [RAGnorm]) and one baseline approach using TF-IDF based String Matching. We evaluated performance across three datasets of SNOMED-mapped condition terms: (1) an oncology-specific dataset, (2) a representative sample of institutional medical conditions, and (3) a dataset of commonly occurring condition codes (>1000 uses) from our institution. We measured performance by recording the mean shortest path length between predicted and true SNOMED CT terms. Additionally, we benchmarked our models against the TAC 2017 drug label annotations, which normalizes terms to the Medical Dictionary for Regulatory Activities (MedDRA) Preferred Terms.

**Results:** We found that RAGnorm was the most effective throughout each dataset, achieving a mean shortest path length of 0.21 for the domain-specific dataset, 0.58 for the sampled dataset, and 0.90 for the top terms dataset. It achieved a micro F1 score of 88.01 on task 4 of the TAC2017 conference, surpassing all other models without viewing the provided training data.

**Conclusion:** We find that retrieval-focused approaches overcome traditional LLM limitations for this task. RAGnorm and related retrieval techniques should be explored further for the normalization of biomedical free text.

## 1. INTRODUCTION

The global transition to digital clinical records has led to a drastic increase in electronically stored unstructured data, making up around 80%(1) of the information within healthcare. This data has the potential to improve patient care, deepen our understanding of diseases, and support research. Yet the different ways medical terms are used in these records make it difficult to extract meaningful insights.(2) For instance, clinical notes often use ambiguous shorthand, inconsistent structure, and domain-specific vocabularies.(3)

Text normalization is the mapping of natural language expressions of concepts into a standardized vocabulary.(4) By creating consistent representations of terms, text normalization helps transform fragmented medical records into structured data that can be more easily analyzed and interpreted. Traditionally, methods have relied upon some form of string matching. These systems follow strict patterns, which can misinterpret or oversimplify clinical language.(5) This results in lost critical information and reduces data reliability for clinical decisions and research.

Semantic embedding methods such as word2vec(6) can transform words into numerical representations, capturing their contextual meanings.(7) This conversion enables computers to understand and process natural language by identifying semantic relationships between words, even when their usage varies. Quantifiable measures such as cosine similarity are used to return mapped terms that are semantically similar to the natural language term.(8) While these methods outperform traditional string matching by capturing meaning beyond exact matches, there are issues when the semantic space becomes denser. When the number of terms in the dataset increases, embeddings may struggle to fully differentiate context, making it harder to identify the most contextually appropriate match. As a result, the top-ranked term returned by the model may not always reflect the intended meaning or context of the input.

Building on the foundation of semantic embeddings, sentence transformers such as BERT(9), BigBird(10), and GatorTron(11) have emerged as state-of-the-art(12)(13) models for Named Entity Recognition (NER) and text normalization. These models extend the capabilities of word2vec by providing deeper contextual understanding and improved semantic differentiation. However, for the purpose of term normalization, it is often necessary to fine-tune these models on specific datasets to ensure their functionality and accuracy in specialized domains.

At the scale at which new generative Large Language Models (LLMs) are now being trained, we can borrow this signal to achieve state-of-the-art performance without the need for fine-tuning. Models like OpenAI’s GPT-4(14), are often deeply pretrained, enabling them to perform well across a wide range of tasks right out of the box.(15) Unlike traditional approaches, modern LLMs can understand nuanced language and making informed predictions based on context, mimicking a human-like reasoning process.(16) In text normalization, this capability enables LLMs to identify and select the most contextually appropriate terms, improving accuracy and consistency when standardizing unstructured medical data.(17)

A recent work by Wang et al.(18) has shown that fine-tuning LLMs with domain-specific corpora can significantly improve their performance in medical text normalization tasks. While effective, this approach is often task-specific and computationally expensive, limiting its scalability and generalizability across different medical domains. Shin et al.(19) found that while fine-tuning can yield high performance, in many cases, prompt engineering a language model can record comparable performance while being more resource efficient. Prompt engineering involves the strategic formulation of input text that leverages the model’s training and capabilities.(20) For instance, instead of using vague or ambiguous language, prompts are crafted to be precise, incorporating specific keywords or phrases that are known to trigger the most accurate responses from the model.

Within prompt engineering, retrieval-augmented generation (RAG) is an approach to enhance model adaptability with minimal added computation.(21) RAG enables systems to extend beyond their inherent knowledge, using external databases to source relevant context when generating text responses. Retrieval-focused methodology has shown promising results in enhancing the quality and applicability of LLM outputs across diverse applications.(22–24)

While these advances in LLM-based approaches are promising, a systematic comparison of different normalization strategies is needed to determine the most effective approach for clinical text. In this study, we develop and evaluate several methodological approaches to text normalization, aligning medical terms with those provided by Systematized Nomenclature of Medicine—Clinical Terms (SNOMED CT).(25) For validation, we tested our approach on Task 4 of the TAC 2017 Adverse Drug Reaction Extraction from Drug Labels Track.(26)

## Statement of Significance

|  |  |
| --- | --- |
| **Problem or Issue** | The challenge of standardizing unstructured medical text in electronic health records (EHRs) for predictive modeling, clinical decision support, and data mining. |
| **What is Already Known** | Traditional text normalization methods rely on string matching, which can misinterpret clinical language, while semantic embedding methods struggle with dense semantic spaces. |
| **What this Paper Adds** | This paper introduces and evaluates LLM-based normalization strategies, particularly highlighting the effectiveness of retrieval-augmented generation (RAGnorm) in improving accuracy and consistency. |
| **Who would benefit from the new knowledge in this paper** | Researchers and practitioners in healthcare informatics, clinical decision support systems, and data mining who want to enhance the reliability and utility of EHR data. |

## 2. METHODS

### 2.1 Synonym Dataset Descriptions

To evaluate the performance of our text normalization methods, we generated multiple datasets of medical terminology based upon the Systematized Nomenclature of Medicine Clinical Terms (SNOMED CT), a fundamental clinical healthcare terminology comprising concepts with distinct meanings and formal logic-based definitions, systematically arranged in hierarchies. We started with a subset of used at our institution, represented as *T* = {*t*_1_,*t*_2_,…, *t*_n_}. For each term *t_i_* ε *T_i_*, we generated a corresponding set of 10 synonyms *Si* = {*s_i_*_1_,*s_i_*_2_,…, *s_i_*_10_} where each synonym *s_ij_* was derived using GPT-4 (version=gpt-4-0125-preview):

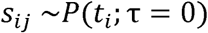

where *P*(*t_i_*; τ = 0) represents the output distribution of GPT-4 conditioned on *t_i_*, with the temperature hyperparameter τ set to 0 to ensure deterministic outputs. A set of example terms are listed in Supplemental Table 1. We assessed our approaches on three different datasets of varying sizes and characteristics to comprehensively evaluate its effectiveness.

#### 2.1.1 Domain Specific Dataset

For the domain-specific data, we concentrated on oncology terminology. We selected terms from the OMOP (Observational Medical Outcomes Partnership) database at our institution, focusing on SNOMED CT descendants of “Malignant neoplastic disease.” Specifically, we identified 106 terms that not only belong to this category but also appear over 1,000 times in our facility’s records.

#### 2.1.2 Cross-Domain: Randomly Sampled

To assess the generalizability of our text normalization methods, we created a cross-domain dataset by randomly sampling 750 terms from a pool of approximately 40,000 terms present in our institutional billing codes. This random selection process ensures a diverse mix of non-domain-specific terms, allowing us to test the adaptability of our models across various medical contexts.

#### 2.1.3 Cross-Domain: Over 1000 Uses

For evaluating the robustness of our models with frequently used medical terminology, we compiled a dataset of 4,747 terms that appear over 1,000 times in our institution’s billing records. This dataset challenges the models with a large volume of commonly used terms, providing a comprehensive test of their capacity to handle extensive and varied medical vocabularies.

#### 2.1.4 Synonym Match Evaluation Metrics

To quantify our results, we take advantage of the domain’s hierarchical nature and use mean shortest path length as the primary metric. This measures the average semantic distance between predicted and true terms within the SNOMED CT ontology. The mean shortest path length is calculated as follows:

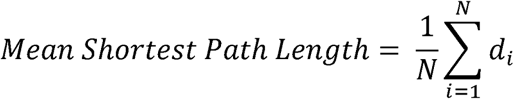

where *N* is the total number of synonyms and *d_i_* is the shortest path length from the *i*th synonym to the correct SNOMED CT.

To give a measure of spread, we used bootstrapping to generate a distribution for the mean shortest path time. This involved resampling the dataset with replacement and recalculating the mean shortest path length across multiple iterations, allowing us to observe variability and derive confidence intervals.

In addition, we monitored the number of API calls made for each method, serving as an indicator of computational efficiency. We also considered the average number of tokens per API call, reflecting the efficiency and cost of communication with the API, as API pricing is based on the number of tokens processed in each call.

### 2.2 TAC2017 Dataset Description

To validate our results on an external dataset, we used the TAC2017 Adverse Drug Reaction (ADR) dataset.(26) This dataset includes a normalization task for adverse event terms mentioned on drug labels to the standardized Medical Dictionary for Regulatory Activities (MedDRA) vocabulary. The dataset includes a set of 200 annotated drug labels with identified adverse reaction terms and their MedDRA mappings. This task is in line with our study’s goal of evaluating text normalization methods, as it necessitates the precise mapping of varied linguistic expressions to a standardized medical terminology system. We used MedDRA v18.1 and augmented our normalization process with the UMLS Terminology Services to assist in mapping the extracted terms to their corresponding MedDRA Preferred Terms (PTs).

#### 2.2.1 TAC2017 Evaluation Metrics

In the evaluation for TAC2017 Task 4, the task coordinators looked at three key metrics: Precision, Recall, and F1 Score, each calculated at both the micro and macro levels. Micro metrics aggregate true positives, false positives, and false negatives across all classes, treating each instance equally, beneficial for datasets with class imbalances. Macro metrics calculate the metric for each class individually and then average them, treating each class equally regardless of size.

### 2.3 Approaches

We present four frameworks to evaluate the capability of LLMs to normalize text. These approaches are visualized in Figure 1. Additionally, we compare an established string-matching technique to serve as a baseline.

**Figure 1:**
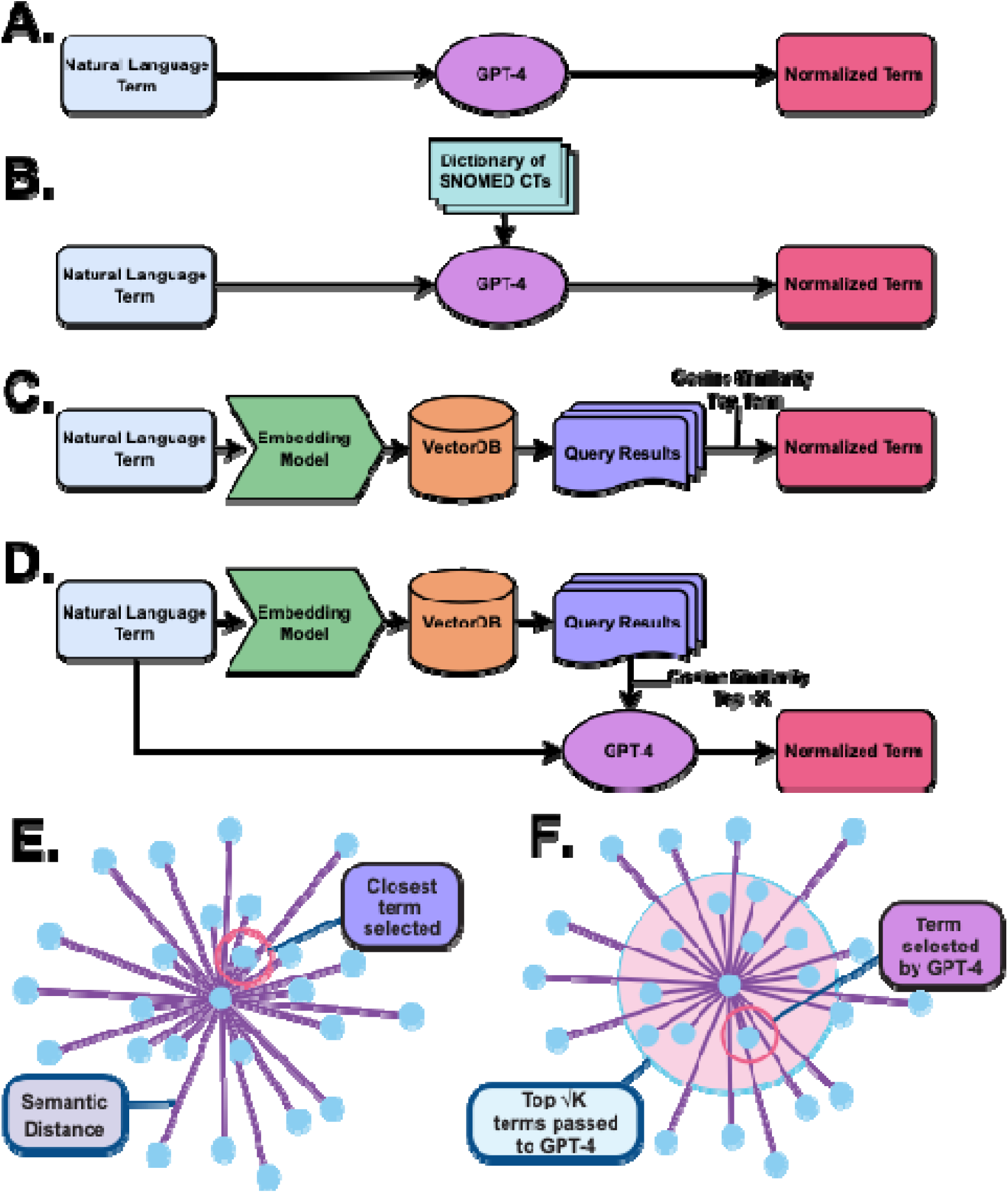
Methodology Flowchart. Step-by-step approach for four different normalization methods— (A) Zero-Shot Recall: Uses a single prompt to elicit the correct term from the model without any prior examples or fine-tuning. (B) Prompt Recall: Feeds the model a comprehensive list of terms, prompting it to select the most appropriate one based on the input context. (C) Semantic Search: Matches input terms with the closest semantic equivalents using a precomputed vector space of embeddings. (D) RAGnorm: First retrieves most semantically relevant terms and then uses the generative decoder to choose the best matching term. An embedding space visualization illustrates the differences between Semantic Search (E) and RAGnorm (F).

#### 2.3.1 TF-IDF String Matching

TF-IDF (Term Frequency-Inverse Document Frequency) String Matching(27) is a method that leverages the statistical properties of terms within a document corpus to determine their importance and relevance. It is calculated by multiplying the term frequency (TF) of a word in a document by its inverse document frequency (IDF) across the entire corpus. This helps in emphasizing terms that are important within a specific document while downplaying common terms that appear frequently across many documents.

In our implementation, each term and synonym are transformed into a TF-IDF vector using the TfidfVectorizer from the sklearn library. To find the closest match for each synonym, we compute the cosine similarity between the TF-IDF vector of the synonym and the TF-IDF vectors of all SNOMED CT entries. The SNOMED CT with the highest cosine similarity is considered the closest match.

This provides a straightforward and interpretable way to normalize text by leveraging the statistical properties of the terms and serves as a baseline for comparing the performance of more advanced techniques.

#### 2.3.2 Zero Shot Prompting

Zero-shot learning(28) is a concept in machine learning where a model makes predictions on data it has never seen during training, effectively requiring it to generalize from previous knowledge without any task-specific fine-tuning. LLMs are trained on diverse objectives that require understanding the relationships between words, sentences, and broader discourse to generate coherent and contextually appropriate text making them especially suited to this strategy. To assess GPT-4’s latent pre-trained knowledge of SNOMED codes, we used a zero-shot prompting strategy. For each term in our datasets, we constructed a prompt that asked the model to provide the SNOMED CT code for a given synonym. (Supplemental Table 2). The model’s responses are then compared to the actual SNOMED CT codes to calculate accuracy. This method was applied to both the domain-specific dataset, which focused on “Malignant neoplastic disease” descendants, and the broader dataset, which included a random sample of terms from commonly used institutional billing codes.

#### 2.3.3 Prompt Recall

In the Prompt Recall approach, we relied on the LLM’s ability to retrieve information from its instruction prompt.(29) For each term in our datasets, we included the entire list of possible SNOMED CT terms and their corresponding IDs (N=106 for the oncology-specific evaluation and N=750 for the broader dataset). This dictionary served as a reference for GPT-4 to accurately recall and provide the SNOMED CT IDs when prompted with the generated synonym. Note that we excluded the high frequency dataset (N=5,300) from this analysis due to prohibitive computational cost (Supplemental Table 3).

#### 2.3.4 Semantic Embedding

Semantic search is a method that leverages the semantic similarity between phrases to improve the accuracy of text normalization.(30) This approach involves using text embedding models to project both the medical terms and the standardized vocabulary into a high-dimensional space where semantically similar terms are closer together. We evaluated embedding models from MTEB Leaderboard(31) and used within the Sentence Transformers Python module. The models under consideration were “text-embedding-ada-002”, “all-MiniLM-L6-v2”,(32) “bge-large-en-v1.5”,(33) “ember-v1”, “GIST-large-Embedding-v0”,(34) “sf_model_e5”, and “SFR-Embedding-Mistral”.(35)

We compared performance of these models against our SNOMED CT reference set (N=750) using the metrics described above. We selected the model that provided the highest accuracy in mapping synonyms to the correct SNOMED CT codes, which we then used for the normalization process in the subsequent phases of our study (i.e. here and for RAGnorm).

#### 2.3.5 Retrieval-Augmented Generation based Normalization (RAGnorm)

Retrieval-Augmented Generation (RAG)(21) combines semantic search and generative language models to perform text normalization. We used a two-step process. First, we used a retrieval system built on GIST-large-Embedding-v0 to fetch the most relevant (based on cosine similarity) SNOMED CT entries based on the input medical term. The top retrieved entries were then passed to GPT-4, which generated the final normalized term by synthesizing the information from the retrieved entries and its pre-existing knowledge. We experimented with several ways to tune the number of entries we retrieve, and ultimately found that taking the square root of the total number of terms gives a manageable yet comprehensive set of results for the language model to work with. (Supplemental Figure 1)

## 3. RESULTS

### 3.1 Selecting an Embedding Model

For each embedding model, we calculated the accuracy through the percentage of synonyms correctly matched to the SNOMED CT codes. The only closed-source model evaluated, “text-embedding-ada-002,” has a dimensionality of 1536 and achieved an accuracy of 74.82%. The smallest model, “all-MiniLM-L6-v2,” uses 384 dimensions and reached 64.73%. Out of the “medium” 1024 dimensional models evaluated, “bge-large-en-v1.5” had an accuracy of 73.59%, “ember-v1” scored 73.34%, “GIST-large-Embedding-v0” produced 76.23%, and “sf_model_e5” achieved 76.08%. The largest model assessed, “SFR-Embedding-Mistral” with a dimensionality of 4096, yielded an accuracy of 81.63%. We chose the “GIST-large-Embedding-v0” for its balance of size and performance for use in the subsequent phases of our study (Figure 2).

**Figure 2:**
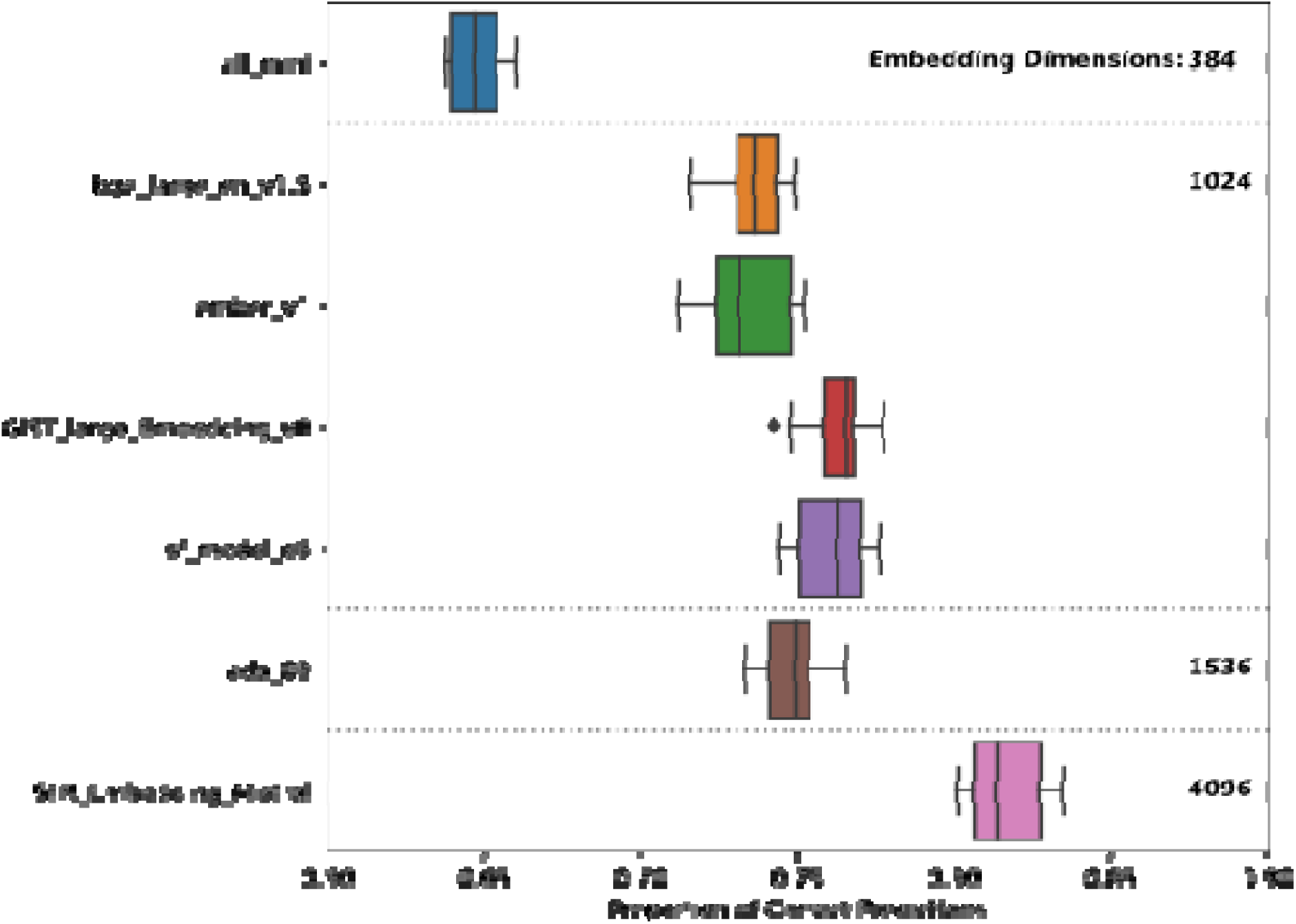
Embedding Evaluation. Comparison of Semantic Search text normalization accuracy across each assessed embedding model. The models are grouped by the number of embedding dimensions used.

### 3.2 Tuning the Number of Terms Returned

RAGnorm required optimization of the number of terms retrieved to inform the language model. We experimented with varying retrieval sizes to find a balance between excluding the true answer and passing a cumbersome amount of information to GPT-4. We identified an optimal number that equated to the square root of the total number of potential terms (Supplemental Figure 1).

### 3.3 SNOMED CT Dataset Results

The TF-IDF-based String Matching gave mean shortest path lengths of 1.02 for the domain-specific set of 106 terms, 1.98 for the randomly sampled set of 750 terms, and 2.37 for the high-frequency set of 4747 terms. The Zero-Shot Recall method recorded mean shortest path lengths of 3.11, 3.80, and 3.80 for the respective cohorts. The Prompt Recall method resulted in mean shortest path lengths of 0.24 and 2.11 for the domain-specific and randomly sampled cohorts, with no data available for the high-frequency cohort. The Semantic Search approach produced mean shortest path lengths of 0.51, 1.25, and 1.17. Finally, RAGnorm attained mean shortest path lengths of 0.21, 0.58, and 0.90 for the domain-specific, randomly sampled, and high-frequency cohorts, respectively. The results of these text normalization methods are presented in Table 1 and Figure 3, with further results and computational complexity metrics reported in Supplemental Table 3.

**Figure 3:**
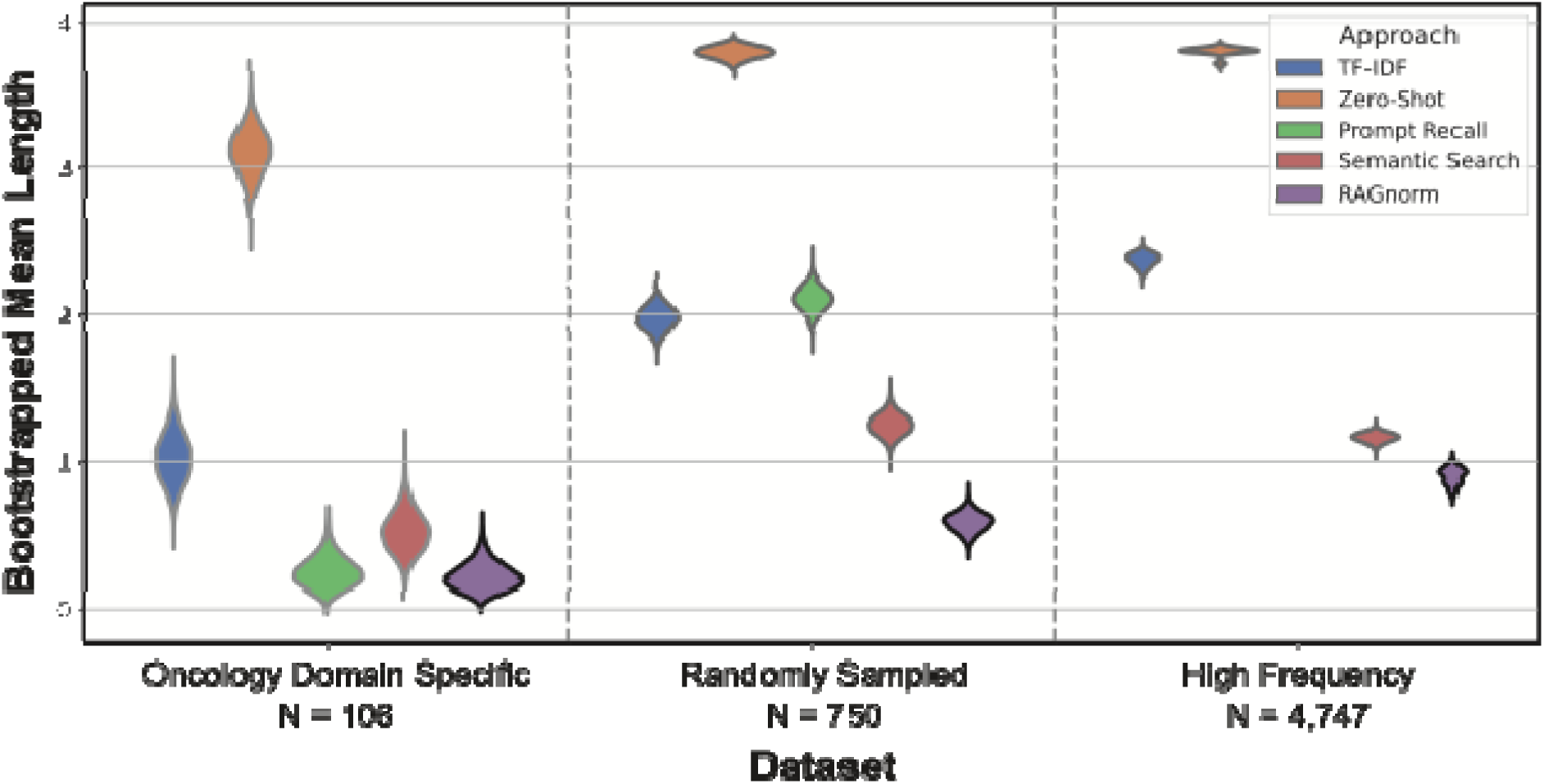
Violin plot of mean shortest path length of five different normalization methods—TF-IDF String Matching, Zero Shot Prompting, Prompt Recall, Semantic Search, and RAGnorm—across three cohort sizes: 106 Domain Specific SNOMED CTs, 750 Randomly Sampled SNOMED CTs, and High Frequency SNOMED CTs (above 1000 uses). The best performing approach on in each dataset grouping is bolded.

**Table 1:** Mean Shortest Path Length Between Predicted and True SNOMED CT Concepts with 95% Confidence Intervals.

|  | TF-IDF | Zero-Shot | Prompt Recall | Semantic Search | RAGnorm |
| --- | --- | --- | --- | --- | --- |
| **Doman Specific**<br>**N=106** | 1.02<br>[0.71–1.37] | 3.11<br>[2.79–3.41] | 0.24<br>[0.08–0.46] | 0.51<br>[0.26–0.80] | **0.21**<br>[0.07–0.42] |
| **Randomly**<br>**Sampled**<br>**N=750** | 1.98<br>[1.83–2.14] | 3.80<br>[3.71–3.87] | 2.11<br>[1.94–2.27] | 1.25<br>[1.11–1.40] | **0.58**<br>[0.45–0.71] |
| **High Frequency**<br>**N=4747** | 2.37<br>[2.28–2.45] | 3.80<br>[3.71–3.85] | --- | 1.17<br>[1.10–1.22] | **0.90**<br>[0.77–1.00] |

### 3.4 TAC2017 Dataset Results

The TAC2017 Adverse Drug Reaction dataset served as a validation set for our normalization methods. Applying RAGnorm to this dataset resulted in a Micro-Precision of 89.6%, Micro-Recall of 86.5%, and a Micro-F1 Score of 88.0%. The Macro-Precision was 89.1%, with a Macro-Recall of 86.1% and a Macro-F1 Score of 87.6%. Our Semantic Search method resulted in a Micro-F1 of 80.83% and a Macro-F1 of 80.55%.

Precision and recall are not included in the table for the semantic search approach because every term is assigned a top-ranked match, leaving no explicit ‘unmapped’ category. In this setup, incorrect matches inherently act as both false positives and false negatives, making precision and recall redundant metrics that are directly reflected in the F1 score. These metrics are tabulated in Table 2, which also includes the performance of other competitors in the TAC2017 (26) challenge, offering a benchmark for RAGnorm’s performance.

**Table 2:** Comparison of our RAGnorm and Semantic Search Framework with TAC2017 Task 4 Results.

| System | Micro-Precision | Micro-Recall | Micro-F1 | Macro-Precision | Macro-Recall | Macro-F1 |
| --- | --- | --- | --- | --- | --- | --- |
| **RAGnorm** | **89.56** | 86.52 | **88.01** | **89.07** | 86.12 | **87.55** |
| **UTH CCB** | 84.17 | **89.84** | 86.91 | 83.02 | **89.06** | 85.33 |
| **UTH CCB** | 85 | 87.75 | 86.35 | 84.04 | 86.67 | 84.79 |
| **UTH CCB** | 82.42 | 90.78 | 86.4 | 80.83 | 89.9 | 84.53 |
| **Semantic Search** | - | - | 80.83 | - | - | 80.55 |
| **CONDL** | 88.81 | 77.16 | 82.58 | 88.2 | 75.76 | 80.5 |
| **PRNA SUNY** | 86.14 | 74.89 | 80.12 | 85.32 | 72.76 | 77.97 |
| **PRNA SUNY** | 81.55 | 78.24 | 79.86 | 79.8 | 76.03 | 77.25 |
| **PRNA SUNY** | 83.6 | 74.14 | 78.59 | 82.22 | 71.44 | 75.87 |
| **CONDL** | 74.56 | 80.96 | 77.63 | 73.06 | 79.92 | 75.55 |
| **CONDL** | 74.56 | 80.96 | 77.63 | 73.06 | 79.92 | 75.55 |
| **MC UC3M** | 73.4 | 80.25 | 76.67 | 72.1 | 80.38 | 75.29 |
| **MC UC3M** | 73.4 | 80.25 | 76.67 | 72.1 | 80.38 | 75.29 |
| **CHOP** | 71.78 | 50.14 | 59.04 | 70.12 | 49.84 | 57.27 |

## 4. DISCUSSION

Our study demonstrates the potential of LLMs to improve the accuracy of medical text normalization, which is an advancement over traditional string-matching or simple machine learning approaches that often struggle with the nuances of medical language.

The Zero-Shot method has fast response times and low computational overhead due to OpenAI’s processing and can be considered a widely accessible, easy-to-set-up option. However, consistent with previous findings,(18) its reliance on specific assumptions about GPT-4’s knowledge of SNOMED CT and its ability to recall details from its training dataset severely limits its accuracy.

The Prompt Recall method benefits from having all the necessary information to select the optimal term. However, this method is hurt by its high token requirements, which can quickly become costly and result in longer compute times, and the performance suffers significantly when given a larger possible set of terms to normalize. This latter limitation, especially, would limit its practical use in real-world scenarios. This limitation was particularly evident when considering the “Cross-Domain High Frequency Terms” dataset, consisting of 4,747 terms. The method requires each potential term to be included in the prompt, making it impractical to apply this approach to such a large dataset due to prohibitive computational costs. As detailed in Supplemental Table 3, the expense associated with processing such a large volume of data within a single prompt was unsustainable, leading us to exclude this dataset from the Prompt Recall analysis.

In contrast, the Semantic Search approach stands out for its speed and cost-effectiveness, leveraging precomputed embeddings to bypass the need for real-time response generation. While its simpler architecture may overlook complex contextual signals, potentially limiting its effectiveness, it strikes a strong balance between efficiency and performance that make it the most desirable option in scenarios where access to a generative LLM is not viable.

Of the approaches, RAGnorm demonstrated the strongest performance in our study. This proficiency can be largely attributed to its dual-phase process, which combines the precision of semantic retrieval with the contextual fluidity of generative language models. RAGnorm’s initial retrieval phase acts as a targeted filter, narrowing down the vast expanse of medical terminology to the most pertinent SNOMED CT entries. This pre-processing step is important, as it primes the model with a distilled set of data that is highly relevant to the input query, thereby increasing the likelihood of accurate normalization. The generative phase of RAGnorm then capitalizes on this curated subset, building upon the retrieved information to produce a normalized output that is not only precise but also contextually enriched. This is particularly helpful when disambiguating terms that have multiple SNOMED CT codes depending on the context.

Methods that rely on prompting often include a large and comprehensive amount of information within the prompt itself, which can be prohibitively expensive in terms of token usage and computational power.(36) In contrast, RAGnorm’s retrieval phase efficiently identifies and retrieves only the most relevant entries, greatly reducing the number of tokens processed (Table 1). This selective retrieval not only saves on computational resources but also streamlines the normalization process, making it more agile and cost-effective.

While more cost effective than other approaches, it is still important to note that requiring both retrieval and generation steps necessitates computational power. The demands translate into potential costs and longer processing times than a simple string-matching approach, which could be a barrier to real-time application in clinical settings. A potential consideration when scaling RAGnorm, adjusting the size of the retrieval dictionary is key. By limiting the dictionary to the most frequently used or contextually relevant terms, we can decrease the number of tokens processed, thus saving computational resources and streamlining the normalization process.

Ultimately, the best approach for text normalization depends on circumstances such as budget, computational resources, and the nature of the target terminology. For instance, well-known concepts like ICD-10 codes may be more suitable for zero-shot prompting since they may be better represented in the LLM’s training data.

As the number of terms in the dataset increased, we observed a consistent decline in the performance of all models. This trend highlights a fundamental problem in text normalization: the complexity of accurately mapping terms grows with the size of the target vocabulary. When potential targets are added, the models face increased difficulty in distinguishing between semantically similar terms, leading to a higher likelihood of incorrect mappings. As the number of potential matches increases, the semantic space becomes more crowded, making it challenging to identify the most contextually appropriate match. This is particularly evident in the high frequency terms (n = 4747 terms) dataset, where the volume of terms leads to a decrease across the board.

The results of our study pave the way for several future research directions. One such direction is the integration of RAGnorm into clinical systems. For example, within Electronic Health Records (EHRs), this could enhance the normalization process by enabling the identification of adverse drug reactions (ADRs) in clinical notes—an area often overlooked by the structured data elements of EHRs. Additionally, there is an opportunity to fine-tune RAGnorm approach for specific medical subdomains, such as genetics or rare diseases, where the terminology is particularly specialized. The incorporation of a user feedback loop could additionally improve our pipeline, where clinicians can provide input to correct and refine the model’s outputs. This could lead to continuous improvement in the model’s performance. Lastly, exploring ways to scale RAGnorm in a cost-effective manner, perhaps by utilizing more efficient language models or distributed computing resources, would make this technology more accessible and practical for widespread use.

This study did not require Institutional Review Board (IRB) approval as it exclusively utilized aggregate data, ensuring that no individual patient information was accessed or analyzed. Furthermore, all interactions with GPT-4 were conducted through a PHI-compliant Azure pipeline, adhering to stringent data privacy and security standards.

## Data Availability

All data produced in the present study are available upon reasonable request to the authors

## ACKNOWLEDGEMENTS

NPT is supported by NIH NIGMS R35GM131905

