## Supplemental Tables and Figures for "Biomedical Text Normalization through Generative Modeling"

Supplemental Table 1: Head of Concept Synonyms

| Concept_Name | Synonym 1 | … | Synonym 10 |
| --- | --- | --- | --- |
| Primary malignant neoplasm of female breast | Breast cancer | … | Malignant breast tumor |
| Primary malignant neoplasm of prostate | Prostate cancer | … | Prostatic carcinoma |
| Primary malignant neoplasm | Cancerous tumor | … | Malignant cancer |
| Primary malignant neoplasm of skin | Skin cancer | … | Cutaneous malignancy |
| Malignant neoplastic disease | Cancer | … | Carcinoma |
| Secondary malignant neoplasm of bone | Bone metastases | … | Metastatic bone cancer |
| Primary malignant neoplasm of respiratory tract | Respiratory tract cancer | … | Respiratory system cancer |
| Secondary malignant neoplasm of liver | Liver metastasis | … | Hepatic metastasis |
| Primary malignant neoplasm of colon | Colon cancer | … | Colorectal cancer |
| Multiple myeloma | Plasma cell myeloma | … | Kahler's disease |

Supplemental Table 2: Model Input

|  | System Prompt | User Input |
| --- | --- | --- |
| Prompt for Synonym Generation | Provide exactly 10 synonyms that are identical in meaning to the given term. Separate each synonym with a '\|' and return no other text besides the 10 synonyms. | SNOMED CT |
| Prompt For Zero-Shot Recall | Provide the most relevant SNOMED CT ID for the given terminology. Return no text beyond the integer ID. | Natural language term |
| Prompt for Prompt Recall / RAGnorm | Provide the most relevant SNOMED CT ID for the given terminology. Return no text beyond the integer ID. Use the following list as a reference: [LIST OF TERMS] | Natural language term |

Supplemental Table 3: Model Performance and Computational Cost

|  | **Domain Specific Terms (N=106)** | | | **Cross-Domain Sampled Terms  (N=750)** | | | **Cross-Domain High Frequency Terms  (N=4747)** | | |
| --- | --- | --- | --- | --- | --- | --- | --- | --- | --- |
| **Approach** | **# API Calls** | **Average Tokens per call** | **Accuracy** | **# API Calls** | **Average tokens per call** | **Accuracy** | **# API Calls** | **Average tokens per call** | **Accuracy** |
| **TF-IDF based String Matching** | 0 | - | 67.0% | 0 | - | 46.6% | 0 | - | 27.7% |
| **Zero-Shot Recall** | 106 | 36 | 4.6% | 750 | 36 | 1.0% | 4747 | 36 | 1.3% |
| **Prompt Recall** | 106 | 1606 | 86.4% | 750 | 11,529 | 40.8% | - | - | - |
| **Semantic Search** | 0 | - | 86.0% | 0 | - | 76.2% | 0 | - | 59.0% |
| **RAGnorm** | 106 | 71.25 | 89.8% | 750 | 139.68 | 80.0% | 4747 | 304.70 | 61.6% |

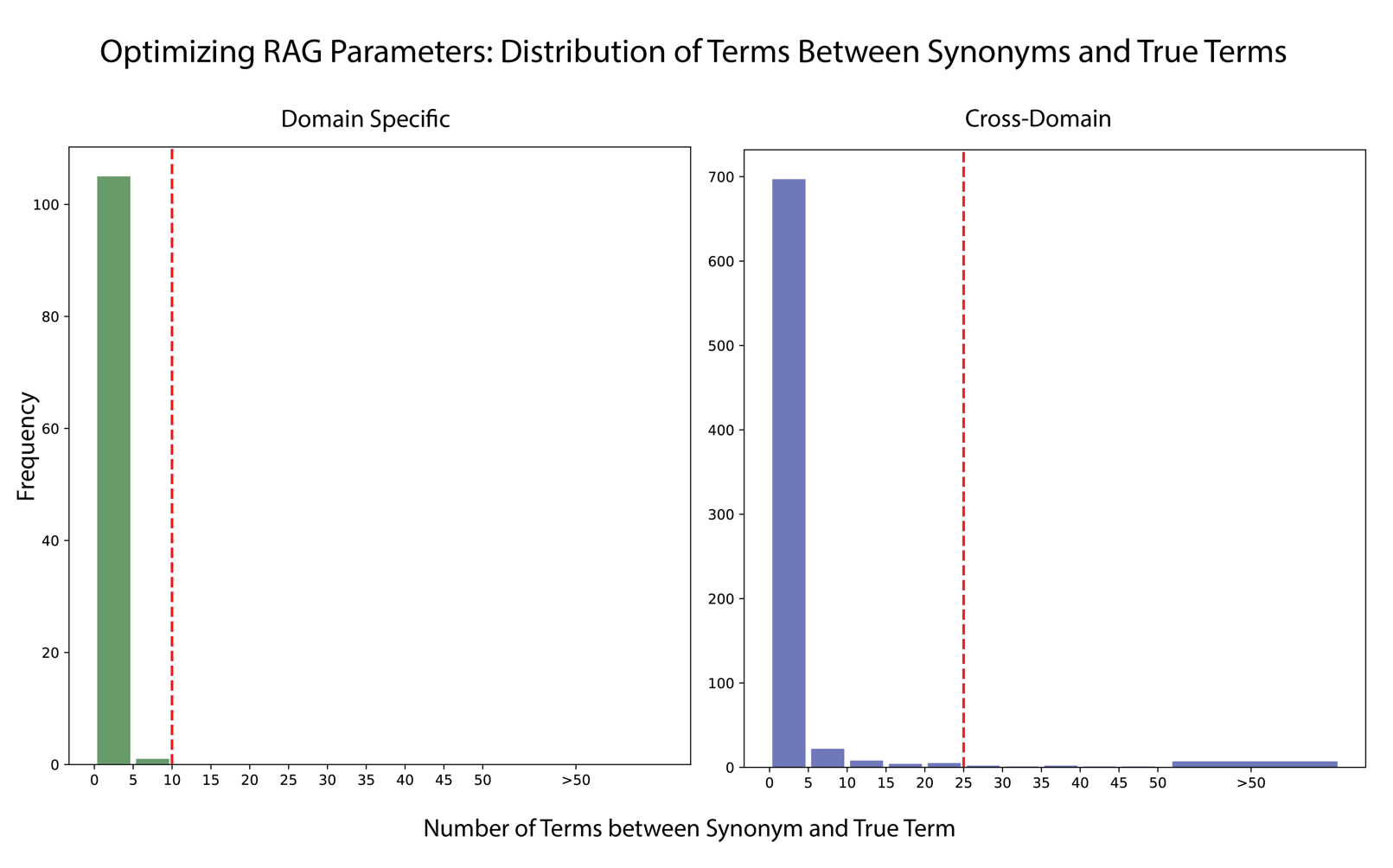

Supplemental Figure 1: Histogram comparing the frequency of terms occurring between synonyms and true terms within domain-specific (106 terms) and cross-domain (750 terms) settings in the semantic space created by GIST-large-Embedding-v0. The x-axis represents the number of terms between synonym and true term, segmented into intervals, while the y-axis indicates the frequency of these occurrences. The red dashed line in each graph denotes the proposed cutoff point at √k, where k is the number of terms.
